# Why lockdown? Simplified arithmetic tools for decision-makers, health professionals, journalists and the general public to explore containment options for the novel coronavirus

**DOI:** 10.1101/2020.04.15.20066845

**Authors:** Gerry F. Killeen, Samson S Kiware

## Abstract

Half the world’s population is already under lock-down and the remainder will have to follow if the ongoing novel coronavirus 2019 (COVID-19) virus pandemic is to be contained. Faced with such brutally difficult decisions, it is essential that as many people as possible understand (1) why lock-down interventions represent the only realistic way for individual countries to contain their national-level epidemics before they turn into public health catastrophes, (2) why these need to be implemented so early, so aggressively and for such extended periods, and (3) why international co-operation to conditionally re-open trade and travel between countries that have successfully eliminated local transmission represents the only way to contain the pandemic at global level. Here we present simplified arithmetic models of COVID-19 transmission, control and elimination in user-friendly Shiny and Excel formats that allow non-specialists to explore, query, critique and understand the containment decisions facing their country and the world at large. Based on parameter values representative of the United Republic of Tanzania, which is still early enough in its epidemic cycle and response to avert a national catastrophe, national containment and elimination with less than 10 deaths is predicted for highly rigorous lock down within 5 weeks of the first confirmed cases and maintained for 15 weeks. However, elimination may only be sustained if case importation from outside the country is comprehensively contained by isolating for three weeks all incoming travellers, except those from countries certified as COVID-free in the future. Any substantive relaxation of these assumptions, specifically shortening the lock-down period, less rigorous lock-down or imperfect importation containment, may facilitate epidemic re-initiation, resulting in over half a million deaths unless rigorously contained a second time. Removing contact tracing and isolation has minimal impact on successful containment trajectories because high incidence of similar mild symptoms caused by other common pathogens attenuates detection success of COVID-19 testing. Nevertheless, contact tracing is recommended as an invaluable epidemiological surveillance platform for monitoring and characterizing the epidemic, and for understanding the influence of interventions on transmission dynamics.

## Introduction

For all but the most expert specialists, the silent early phase of an epidemic, when there is still time to contain it, is often imperceptible and difficult to grasp the significance of. This is especially true for pathogens like novel coronavirus-2019 (COVID-19) that largely exhibit only mild symptoms, if any,^1-23^ that are non-specific and difficult to distinguish from other, more common illnesses.^24^

Currently, half the world’s population is already under lock-down of some kind, meaning vertically enforced and severe restrictions of movement, and the remainder will have to follow if the ongoing COVID-19 virus pandemic is to be contained. Faced with such brutally difficult decisions, it is essential for policy-makers, health professionals, journalists and the general public that as many people as possible understand (1) why lock-down interventions represent the only realistic way for individual countries to contain their national-level epidemics before they turn into public health catastrophes, (2) why these need to be implemented so early, so aggressively and for such extended periods, and (3) why international co-operation to conditionally re-open trade and travel between countries that have successfully eliminated local transmission represents the only way to contain the pandemic at global level.

### An educational tool to help non-specialists understand COVID-19 transmission dynamics and containment strategies

Here we introduce a simplified *arithmetic modelling* tool for predicting COVID-19 transmission dynamics and how it is likely to respond to different containment, delay or mitigation strategies. We coin the term *arithmetic* modelling, as distinct from the ubiquitously used term *mathematical* modelling, to convey the fact that it uses only addition, subtraction, multiplication, division, rounding off, a few conditional statements (eg. *if, less than*/*greater than, and/or*), and two unavoidable power terms, to make the necessary calculations. This tool includes no differential equations, calculus, limits, distributions, stochastic simulations or agent-based approaches that would render it opaque to most non-specialist readers, such as medical and public health practitioners, decision-makers, journalists and the general public. The model is presented in user-friendly Excel^®^ and Shiny^®^ formats that allow non-specialists to explore, query, critique and understand the containment decisions facing their country and the world at large. For those who wish to satisfy themselves that the calculations make intuitive sense, the Excel^®^ version provides a complementary spreadsheet format in which the formula for each cell can be critically examined. For those content to accept the underlying arithmetic, the Shiny^®^ format provides a convenient interactive web application that can be used on any device. While a formal mathematical description of this model has been critically reviewed by specialist experts, it is provided only as an online supplement because none of the principles, assumptions or predictions are entirely new,^25-50^ and because in our experience nothing deters non-specialists from reading an article faster than equations do.

We caution readers not to expect too much from any predictive model in terms of exact numerical reliability,^51-53^ and note that this one is no different. We specifically advise against interpreting the exact numbers this tool generates at face value: Any predictive model is, by definition, a deliberately simplified representation of complex real-world processes, the usefulness of which is largely subjective.^51-53^ The exact numerical predictions should therefore not be used to confidently define precise operational timelines for introducing and sustaining interventions, or set effectiveness thresholds required of specific containment measures. Instead, the purpose of this tool is to help users broadly understand the inevitable consequences of an uncontained epidemic, explore the likely outcomes a wide range of different possible containment strategies, identify those which could plausibly succeed and understand the failures of those which seem unlikely to do so.

Any users finding themselves forced to use numerical predictions from this model to make programmatic intervention decisions, presumably for want of a more reliable alternative in their specific context, should therefore assume sizable imprecisions that will not be possible to quantify prospectively and will be too late to quantify retrospectively. They should therefore factor such unknown levels of uncertainty into their response plans by allowing for wide margins for error when planning the timing, intensity and duration of new interventions, always being more ambitious and cautious whenever in any doubt.

### Tanzania as an illustrative example of national containment options and requirements

Assumed input parameters values were chosen to be representative of the United Republic of Tanzania (Table 1), because it has experienced relatively modest inbound air traffic from China^54,55^ and is still early enough in its epidemic cycle and response for a national catastrophe to be averted. Tanzania has also had more opportunity to learn from ongoing experiences in Asia, Europe and north America, and prepare by establishing testing capacity at the outset of the national epidemic, more consistent with that simplifying assumption of the model than Asian and European countries affected earlier in the pandemic would be. Tanzania is also a typically vulnerable, low-income African country, ^55-60^ which had only 38 ICU beds in all four national referral hospitals combined in 2019,^61^ and is representative of the pandemic that is now imminent all across Africa. ^56,62^

**Table 1.** Assumed values for input parameters of the arithmetic model as intended to be representative of COVID-19 transmission and successful epidemic containment in the United Republic of Tanzania (Figure 1). A detailed formal description of how the model calculations are made, the underlying assumptions are provided in the online methodological supplement to this paper.

| Input parameter description | Assumed value | Dimension | References |
| --- | --- | --- | --- |
| Basic reproductive number (Average number of new infections arising from a single existing infection over its full duration if allowed to do so in a fully susceptible, immunologically naïve population in the absence of any control measures) | 4.0 | Number | 31-33,42,43,50, 63-68 |
| Duration of infection (Average number of weeks an infection lasts in a human before it is eliminated by the immune system). | 3 | Time | 2,3,5,7,15,6 9,70 |
| Human population size | 57 million | Number | 71 |
| Baseline incidence of unrelated similar symptoms (Proportion of population per week experiencing similar symptoms to COVID-19 but caused by other common pathogens like the common cold, influenza, malaria, etc) | 1% | Time<sup>-1</sup> | 24,72 |
| Initial importation rate (Number of new primary cases arriving into the country each week) | 5 | Time<sup>-1</sup> | Assumed |
| Time to initiation of importation containment intervention at border posts, airports and ports of entry (Number of weeks since the first imported cases before inbound travellers to the country are isolated on arrival) | 2 | Time | Assumed |
| Time to initiation of lock-down intervention (Number of weeks since the first imported cases before population-wide restrictions are introduced to prevent personal exposure behaviours) | 5 | Time | Assumed |
| Duration of lock-down intervention (Number of weeks since initiation of population-wide restrictions to prevent personal exposure behaviours until these restrictions are lifted) | 15 | Time | Assumed |
| Asymptomatic proportion of cases (Proportion of all cases who lack, don't notice or don't report any overt symptoms associated with the infection) | 50% | Dimensionless | 8-12,21,25,73,74 |
| Proportion of symptomatic cases which are clinically severe (Percentage of all cases exhibiting and reporting with severe symptoms, all of whom are assumed to be tested unless the limits of testing capacity are exceeded) | 20% | Dimensionless | 8,75 |
| Proportion of mild and severe symptomatic cases requiring critical care (Percentage of all cases exhibiting and reporting with any mild or severe symptoms who need intensive care, which can also be described as $\theta_{m+s,c}$) | 4% | Dimensionless | 8,75,76 |
| Intensive care unit (ICU) capacity (Maximum achievable percentage of the population that could be admitted to an ICU at a given time, allowing for maximum emergency expansion of capacity at short notice) | 0.0002% | Dimensionless | 61 |
| Case fatality rate in ICUs (Percentage of cases needing intensive care who access it but die nevertheless) | 20% | Dimensionless | 75 |
| Case fatality rate outside ICUs (Percentage of cases needing intensive care but who cannot access it and die subsequently) | 50% | Dimensionless | Assumed |
| Maximum achievable diagnostic testing rate (Percentage of entire population per week) | 0.02% | Dimensionless | Assumed |
| Proportional containment of imported cases (Percentage of secondary cases arising from primary imported cases which are prevented by travel restrictions and isolation of inbound travellers from affected countries on arrival) | 100% | Dimensionless | Assumed |
| Proportional containment of contact clusters of confirmed cases (Percentage of secondary cases arising from diagnostic-confirmed primary cases which are prevented by contact tracing and isolation) | 90% | Dimensionless | 24,77 |
| Proportional lock down effectiveness (Percentage reduction of exposure behaviours, eg. close personal contact, sharing venues, transport, goods and other objects, among the fraction of the population included in and compliant with the lock down interventions) | 90% | Dimensionless | 8 |
| Proportional lock down coverage (Percentage of entire population included in and compliant with interventions to reduce exposure behaviours, inclusive of staying indoors, avoiding other people, wearing face masks, and frequent hand washing) | 90% | Dimensionless | 8 |

### COVID-19 may be eliminated and excluded by ambitious national containment campaigns

The simplified model predicts that national containment and elimination may be achieved and sustained, without ever exceeding national ICU capacity, by using a full, timely package of interventions. The national epidemic may be contained with only 1486 cases and 6 deaths by highly rigorous 15-week lockdown (90% effective exposure prevention behaviours by 90% of the population) as soon the first cases are confirmed, 5 weeks into the epidemic, complemented by 90% effective tracing and isolation of all contacts for confirmed cases (Figure 1).

**Figure 1.**
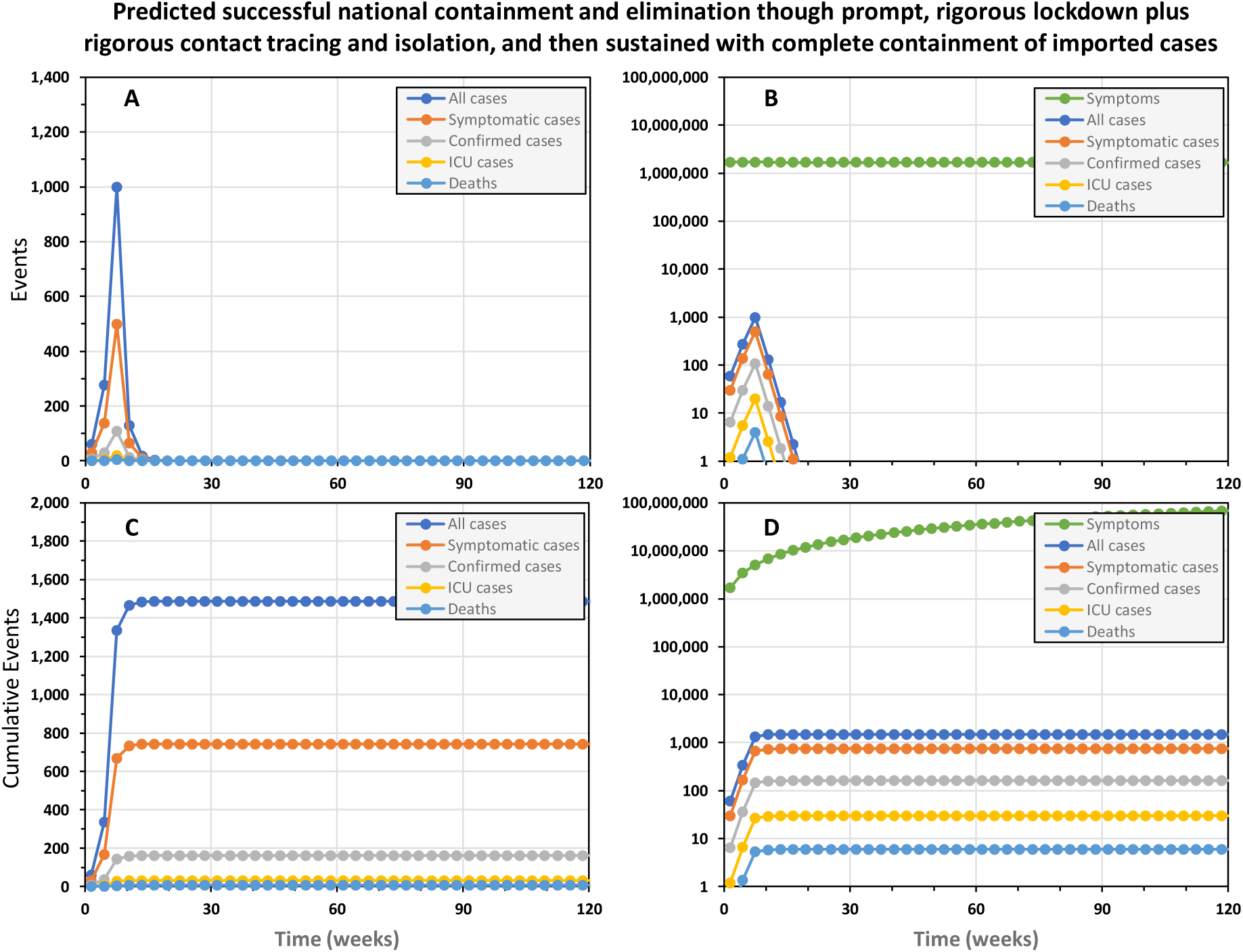
The predicted trajectory of a successfully contained national COVID-19 epidemic in the United Republic of Tanzania. In this simulation, a rigorous 15-week lock down was initiated from week 5 onwards and complemented by complete containment of imported cases, as well as contact tracing and isolation of confirmed cases. Rigorous lock down was assumed to achieve 90% reduction of exposure behaviours by 90% of the population. Complete 100% containment of imported cases assumes that all inbound international visitors are fully isolated for three weeks,^2,3,5,7,69,70,78^ except those coming from countries that may be certified as free of local transmission by WHO in the future. Contact tracing and isolation follow up from confirmed cases was assumed to be 80% effective at preventing onward transmission from entire contact clusters.

As points of reference against which ongoing national containment campaigns may benchmark themselves, the epidemic was predicted to grow 59% bigger each week at the outset and shrink by 48% each week once rigorous lock down had been in place for several weeks. Note, however, that even these alarming projections for the rate of expansion of the epidemic and the rate of contraction required to contain it may under-represent the scale of the challenge in real epidemics. For example, at the outset of the epidemic in China, numbers of confirmed cases doubled every week.^31,79^ Furthermore, subsequent analyses allowing for frequent carriage without overt symptoms indicate much higher viral reproduction rates than assumed in table 1, and suggest true doubling time for all cases may be less than 3 days.^65,67^

Interesting, almost exactly the same containment trajectory is predicted even if contact tracing and isolation is completely removed from the intervention package (Supplementary figure 1), resulting in only 276 more cases and one more death. The explanation for this becomes apparent when one examines the trajectories of confirmed versus all cases: Even though the number of real cases never approaches an optimistically-assumed full testing capacity of 11,400 patients per week, half of all cases are never tested because they are asymptomatic and most of the remainder are only mildly symptomatic, so they get lost in the mass of other people who appear equally sick for unrelated to COVID-19. As illustrated in figure 1D, the background noise of similar mild symptoms caused by other common pathogens dwarfs the mild COID-19 cases, so almost all of them go untested and undetected. Less than one in every 4000 tests is conducted on a mildly symptomatic case of COVID-19, so even though we assume all severe cases are tested, only 11% of cases predicted to occur were confirmed. With contact tracing and isolation only being possible for this very small fraction of cases, there are obvious limits to how much it can achieve as a containment intervention in its own right.

### Even the slightest relaxation of lock down or importation controls cause containment failure

However, successful containment (Figure 1) does requires that the lock down intervention is maintained for the full 15 weeks (Figure 2A and B) to eliminate the virus. Delaying a 15-week lock-down by only 3 weeks, the duration of one generation of viral infection, also allows the virus to persist and the epidemic resumes soon afterwards (Figure 2C and D). A slightly less rigorous lock down of the same duration, which nevertheless achieves 80% coverage with 80% reductions of personal exposure behaviours, also fails to eliminate the epidemic with tragic consequences (Figure 2E and F).

**Figure 2.**
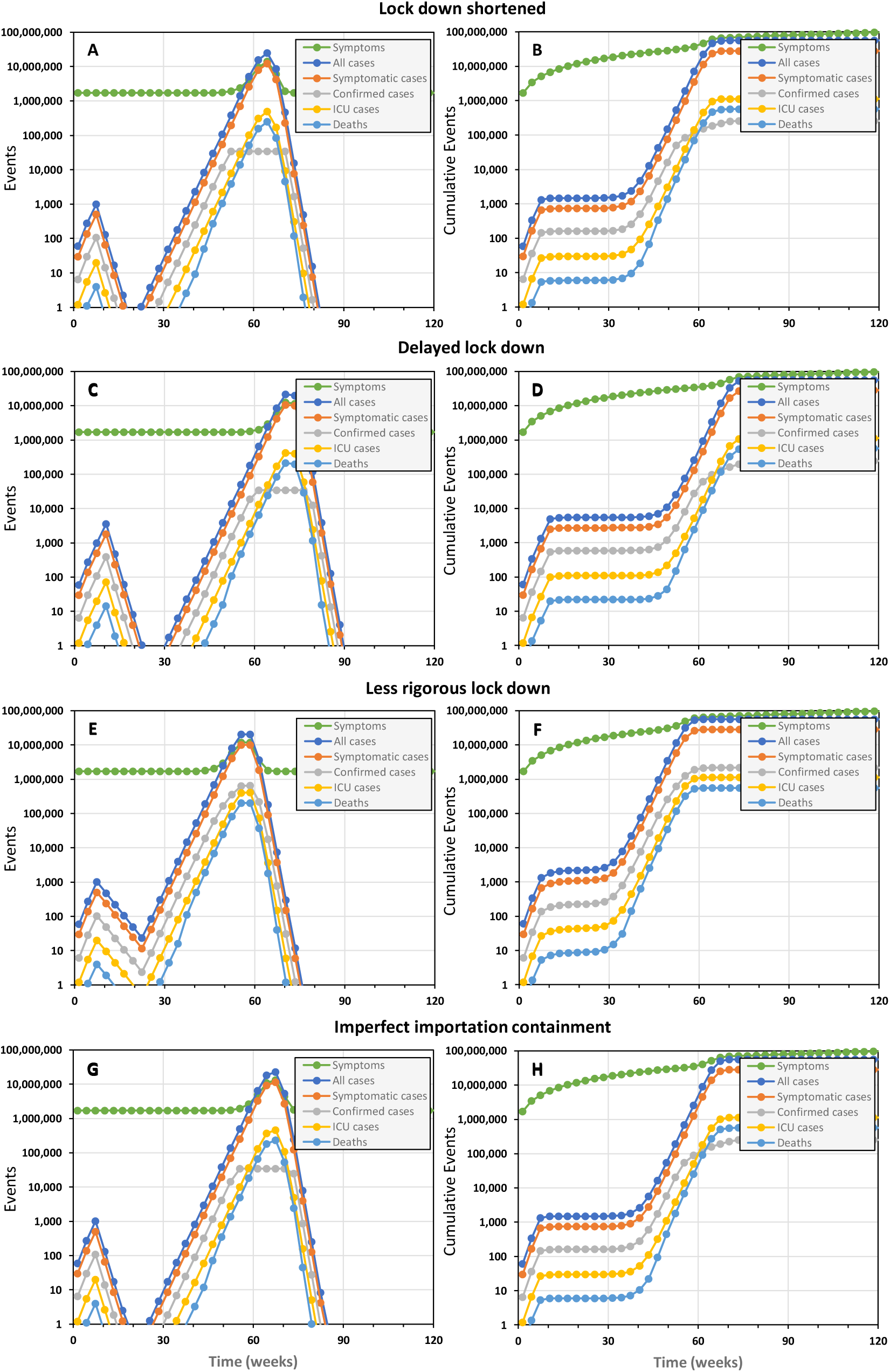
The simulated epidemic trajectories for slightly less robust national COVID-19 epidemic containment responses in the United Republic of Tanzania than that illustrated in figure 1, all of which are predicted to fail and result in a catastrophic rebound of transmission, morbidity and mortality. All these simulations have identical input parameters to figure 1 except for (1) shortening the lock down period by 3 weeks, from 15 to 12 weeks (Panels A and B), (2) reducing importation containment from 100% to 90%, (3) delaying the lock down by 3 weeks (Panels C and D), starting on week 8 rather than week 5 (Panels E and F), and (4) reducing the coverage and protective effectiveness of exposure behaviour reduction from 90% to 80% (Panels G and H).

Furthermore, elimination may only be sustained by comprehensively containing case importation from outside the country (Figure 2G and H). Preventing reintroduction requires isolation of <u>all</u> incoming travellers, except those coming from countries that may be certified as free of local transmission by WHO in the future, to achieve 100% prevention of onward local transmission (Figure 1). Even 90% containment of imported cases seems unlikely to protect the country against reintroduction of the virus and re-initiation of the epidemic (Figure 2G and H). Tanzania therefore did the right thing by isolating all inbound travelers since March 23^rd^ (over 600 so far) for two weeks following their arrival). However, for such importation containment measures to effectively exclude new cases from a COVID-free Tanzania in the future, isolation periods may need to be extended to three weeks.^2,3,5,7,69,70,78^ However, it is also notable that all the scenarios in figure two, except for panels G and H, assume 100% effective containment of imported cases. It is therefore clear that local transmission must be eliminated before such rigorous control of inbound travellers can usefully protect the country against reintroduction.

All these delays, truncations or inadequacies of lock down, or imperfections of importation containment, result in failure to eliminate local transmission that then rebounds and rapidly spirals out of control without a second full containment campaign (Figure 2). The implications of such an uncontained rebound scenario are essentially identical to doing nothing in the first place: In all cases, 99% of the population is expected to become infected over about a year, resulting in approximately 540,000 deaths and ICU demand exceeding capacity about 800 times over. It is also worth noting that total national hospital inpatient capacity of approximately 50,000 beds^80^ would be overwhelmed by cases of severe COID-19 disease peaking at 2.3 million over a three-week period. Under such conditions of a full-blown public health catastrophe, the mitigating effect of stronger health systems in high income countries are largely negated, so our predictions of over half a million deaths in Tanzania compare well with those of others for the United Kingdom,^29^ which has a similar population size. Considering also the travel distances and household costs of hospital attendance in Tanzania^81-84^, it also raises the question as to whether severe COVID patients should be cared for in hospitals and other health facilities^85-88^ which are already 52% understaffed^89^ or at home^85-88^ with support from a rapidly mobilized cadre of Community Health Workers, for which well-characterized curricula and training platforms already exist.^90-92^

### The mirage of *flattening the curve* to steadily acquire population-wide *herd immunity*

The best-case scenario we could identify for “flattening the curve”, as advocated by many national and international authorities, required removing all importation controls to ensure steady re-seeding of the epidemic with a small number of cases and relaxing lock down assumptions to exactly 69% effective reduction of exposure behaviours among 69% of the population (Figure 3C and D). Under such precisely assumed conditions, the epidemic proceeds steadily with between 7 and 9 ICU cases per week over a decade, at the end of which national ICU capacity has never been exceeded and only 1085 deaths will have occurred. However, at the end of such a 10-year campaign, with no end in sight for at least several decades, only 0.5% of the population would have acquired hard-won immunity through prior infection, so the remainder of the population would remain just as vulnerable to a resurgent epidemic.

**Figure 3.**
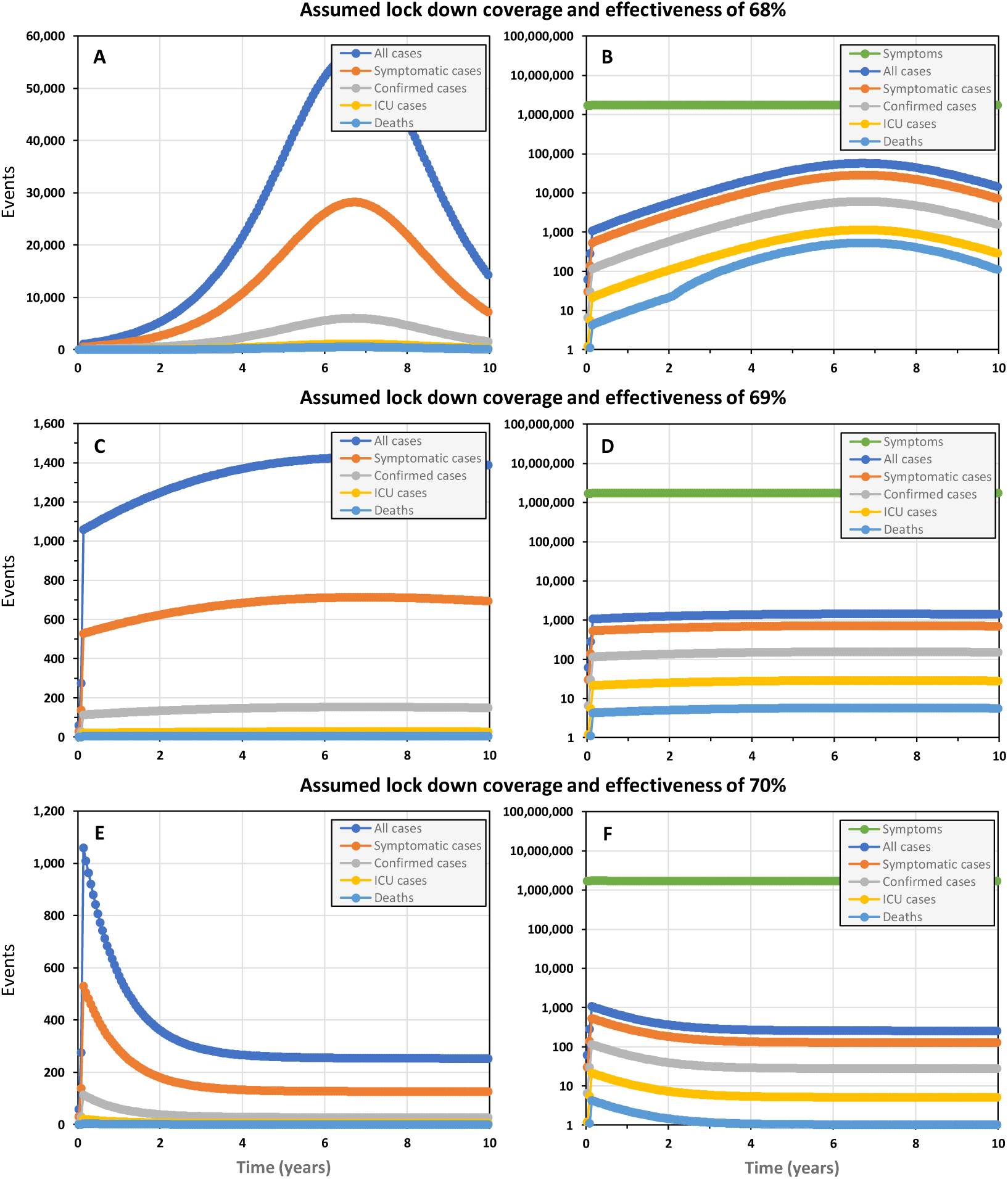
The simulated epidemic trajectories for a less robust national COVID-19 epidemic containment responses in the United Republic of Tanzania than that illustrated in figure 1, intended to *flatten the curve* enough for national health system capacity to cope while *herd immunity* is acquired over the long term. All these simulations have identical input parameters to figure 1 except that no importation containment is assumed and the coverage and protective effectiveness of exposure behaviour reduction is assumed to be lower, at 68% (Panels A and B), 69% (Panels C and D) or 70% (Panels E and F).

However, such precise control over real epidemics with such sensitive and extremely curved trajectories, will be unachievable in practice. Even lowering the assumed lock down coverage and effectiveness parameters for a simulated epidemic by only 1% to 68% results in a long drawn out peak that completely overwhelms ICU capacity within 3 years and continues to do so after a decade (Figure 3A and B), nevertheless leaving 91% of the population lacking acquired immunity. On the other hand, raising assumed lock down coverage and effectiveness by only 1% to 70% results in a long-drawn out containment trajectory that never reaches the elimination end-game (Figure 3E and F) because the steady trickle of imported cases sustains transmission. Re-introducing complete containment of imported cases merely results in a more extended version of Figure 2E and F, with elimination taking over 6 years to achieve (Supplementary figure 2).

Perhaps more to the point, simply expressing ICU capacity as a proportion of overall population size pragmatically puts suggestions that countries should aim to merely slow and mitigate their COVID-19 epidemics into stark perspective. Even if Tanzania can build its ICU capacity from 38 to 114 beds in the coming weeks, and even if the whole population could be somehow perfectly queued up for COVID-19 exposure to make full sequential use of that capacity, assuming each patient needs only 1 week in the ICU and all regular causes of ICU admission magically disappeared, it would take almost two centuries to care for the 1.14 million COVID-19 cases expected. Readjusting such hypothetical calculations to represent higher capacity countries like Ireland or the UK shortens these timeline to decades rather than years, so “flattening the curve” to achieve population-wide “herd immunity” is clearly an infeasible and unwise choice.

### The spiralling costs of catching up on lost time to implement a lock down

If a lock down is delayed by three weeks, approximately the duration of one viral infection, the epidemic may still be contained be extending it by the same length of time, from 15 weeks to 18 weeks (Figure 4A and B). Note, however, that the epidemic peaks at an almost four-fold higher incidence of cases, resulting in 5,485 cases and 22 deaths overall. Although ICU capacity is not expected to be overwhelmed, timely access will clearly represent a challenge for many patients in country with a surface area of almost a million square kilometres and only four national referral hospitals. Longer delays of 6, 9 and 12 weeks necessitate prolonged lock downs (21 weeks for the latter) to contain epidemics of rapidly expanding scale: 19925, 77055 and then 260103 cases, exceeding ICU capacity by 151, 994 and 4597 patients, and resulting in 125, 586 and 2420 fatalities, respectively.

**Figure 4.**
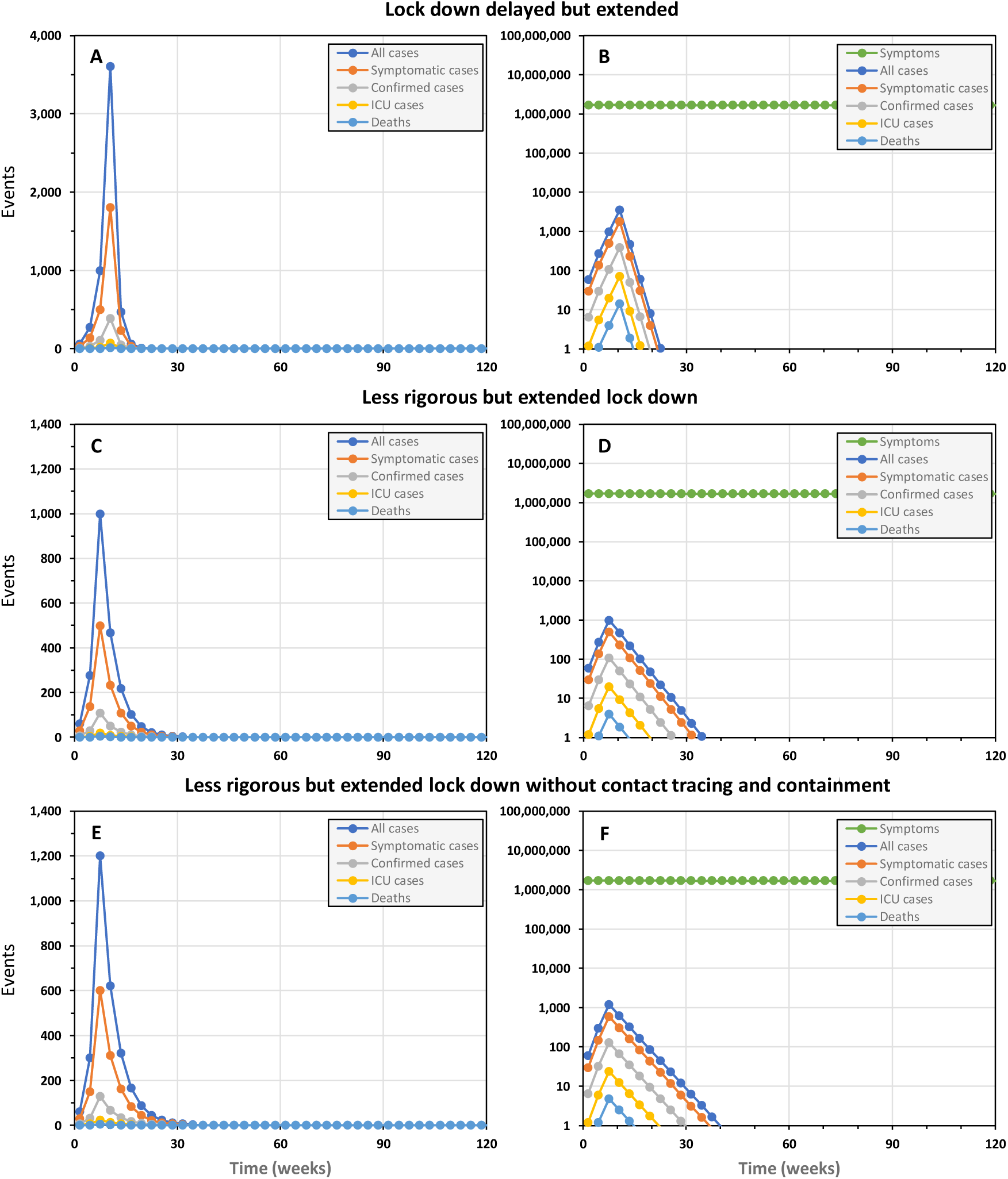
The simulated epidemic trajectories for national COVID-19 epidemic containment responses in the United Republic of Tanzania with slightly delayed or less rigorous lock down than illustrated in figure 1, all of which necessitated extension of the lock down period to achieve successful containment. All these simulations have identical input parameters to figure 1 except for (1) delaying the lock down by 3 weeks, starting on week 8 rather than week 5 (Panels **A** and **B**), (2) reducing the coverage and protective effectiveness of exposure behaviour reduction from 90% to 80% (Panels **C, D, E** and **F**), (3) removing the contact tracing and isolation component (Panels **E** and **F**), and (4) necessarily extending the lock down period from 15 to 18 weeks (Panels **A** and **B**) or from 15 to 40 weeks (Panels **C, D, E** and **F**).

### The hidden dangers of stealthy epidemics

Note, however, that none of this will be obvious during the silent early phase of the epidemic, during which time the number of undetected cases snowballs: Even if the lock down response is initiated after only 5 weeks post-initiation, immediately after the first 6 cases are confirmed in this simulation, the epidemic has already quietly progressed much further than most members of the public would guess. Indeed, far enough that another 271 people are already actively infected and almost 1000 new cases are predicted to occur in the subsequent 3-week period, out of which only 108 (11%) will be detected.

Infectious carriers who exhibit little or no symptoms at the time^1-23^ clearly contribute to the cryptic nature of an early-stage COVID-19 epidemic: In this case we assumed this accounts for 50% of cases lacking symptoms overt enough to consider self-reporting and seeking a test (table 1). However, a much more important factor is the sheer volume of background noise arising from similar symptoms caused by more common pathogens, such as the common cold and malaria. Even though these simulations assume that capacity for conducting 11,400 COVID-19 tests per week would have been established in Tanzania before the outbreak began, total confirmed cases are only expected to exceed 100 about 3 weeks after the lock down is introduced. Most of these confirmed cases are accounted by the clinically severe fraction we assume will all be tested. Only 4% (8/899) of predicted mild or asymptomatic cases are expected to be confirmed because the relatively small number of COVID-19 cases are so easy to miss in a population of 57 million people, out of whom we assume 1% or 570,000 will experience a fever, cough or stomach pains in any given week for unrelated reasons (Figure 1B and D). Note, however, that that even this is a very conservative assumption about background rates of illness with similar symptoms to COVID-19: In the first contact-tracing study in the USA, over 12% of all carefully-followed contacts became symptomatic within 2 weeks, even though none of them became infected with COVID-19.^24^

These simulations are nevertheless useful in that they illustrate how no perceptible increase in the incidence of such common symptoms may be obvious to the general population unless containment efforts fail and a full-scale, resurgent epidemic sweeps through the country (Figure 2). Tanzania therefore did exactly the right thing by reacting fast during the silent earliest phase of the epidemic, announcing school closures within a day of the first confirmed case report and introducing additional restrictions immediately afterwards.

Note, however, that the quiet tail of a fading epidemic may be just as dangerous as the it’s silent onset. Three of the four scenarios in figure 2 include periods of two month or more when few if any confirmed cases are expected, but some mild cases persist that can re-seed the whole epidemic afresh. The predicted persistence of the epidemic despite total predicted cases dropping below zero is an artefact of the simplified deterministic form of the model, which calculates case numbers as a continuous decimal outcome until it drops below 0.1, at which point it is set to 0 because the probability of elimination is 90% or better. The take-home message is nevertheless clear: persist with rigorous lock-down until one can be sure that elimination has been achieved, drawing on statistical approaches used by veterinary epidemiologists to certify elimination with imperfect surveillance systems.^93-98^

### The vital importance of ambition and rigour to lockdown outcome: Who dares loses least!

As illustrated by figure 4C and D, it is now crucial that Tanzania urgently builds on that early momentum to ramp up lock down efforts to the most rigorous level practically attainable. The implications of even a slightly less rigorous lock down appear less daunting in epidemiological terms but far more severe in practical and economic terms, because it greatly prolongs the lockdown period required: Even reducing coverage and effectiveness of personal protection measures by only 10%, from 90% to 80% requires that the lock down period is extended by more than 150%, from 15 weeks to 40 weeks (Figure 4C and D). The practical social and economic sustainability of such a protracted lock down period is very questionable, but much can be learned from the predicted benefits of getting such an imperfect lock down started in good time: The 1,108 cases and 11 deaths predicted over the course of such a “slow burn” containment campaign are only marginally higher than for the best case scenario illustrated in figure 1. It is therefore important to get some form of reasonably rigorous lock down in place as early as possible, and then intensify it as rapidly as possible. Like any race, it is critical to get an early head-start by any means possible, but then build up speed towards a strong finish.

While compliance and enforcement is of great importance to lock down effectiveness, so is acceptability and socio-economic feasibility. While high income countries move to facilitate population-wide compliance with direct financial support and augmented social services, different tactics will be required in low income countries like Tanzania. Although Tanzania is urbanizing very rapidly, most of the population still resides in rural areas^99^ where propagation of directly-transmitted diseases like COVID-19 is always less intense. Fortunately, Tanzania is currently in the midst of this year’s farming season, during which many rural families are out in the fields where social distancing is relatively easy. While farming season also conveniently brings a lull in trading activity at commercial hubs in rural towns and villages, it also represents a seasonal low point in the domestic food reserves of many rural households, so selective food support may be invaluable for enabling the most vulnerable families to comply effectively with self-quarantine and self-isolation directives. However, the growing urban population represents a much larger challenge, because far fewer people rely on farming for their livelihoods. Many live in crowded informal settlements where lack of shelter, water, sanitation and space, relying on unreliable, informal sources of income to survive on a day-to-day basis. Informal livelihoods and settlements in the busiest urban centres of the country will therefore require particularly urgent attention and creativity, to support daily food, water and hygiene needs. It may also be useful to consider providing safe transport with managed social distancing (to be followed by self-isolation) for those with options to sit out the epidemic with family and friends in rural areas.

As is the case for elimination of other diseases, such as malaria for example^100^, it may be more useful to think about gaps in coverage and effectiveness to understand how such apparently minor deficiencies can make all the difference between success and failure: While a shift from 90% to 80% lock down coverage and effectiveness might seems small in relative terms, a 20% shortfall relative to perfect containment is twice as big as 10%. And the difference between 100% prevention of onward local transmission from imported cases contrasts starkly with even such high targets as 90%: when you need to achieve zero new cases in a country, any other number simply isn’t good enough.

In practical terms, we should first think of most the vulnerable, such those lacking homes, shelter, security, citizenship or family support,^101-103^ especially those in the low income countries at greatest risk.^55,57,59,60,101^ Beyond these long-neglected population groups, the most important lock down coverage and effectiveness gaps will be accounted for by the most important exceptions to restrictions and those exceptional individuals most determined to evade them.

Unfortunately, the most obvious exceptions to lock down restrictions who will facilitate continued transmission will be health service personnel,^104-107^ notably those caring for those most vulnerable to the disease. However, all other essential workers in shops, markets, kitchens, food processing facilities, factories, banks, post offices, transport services and law enforcement agencies will also inevitably mediate more transmission than they would if they stayed at home. Indeed, it the crew that enabled self-sustaining levels of COVID-19 transmission to persist aboard the quarantined Diamond Princess cruise ship.^108^ It is also worth remembering that the anti-hero of infectious disease epidemiology, the infamous *Typhoid Mary* (real name Mary Mallon) was a cook by profession who infected at least 53 people, three of whom died.^109^ Like many COVID-19 cases,^1-23^ Mary was a silent carrier of the disease: She herself lived to a ripe old age and died of a stroke rather than typhoid. This is not to say that such essential services should necessarily be suspended, but rather that the roles and working practices of these personnel should be scrutinized particularly carefully. How essential is essential? What is the minimum level of service needed to facilitate extended lock down while mitigating indirect effects on health, well-being and economic welfare that are even worse than COVID-19? What procedures, behaviours and protective equipment could most effectively minimize persistent workplace transmission?

And with so many people’s livelihoods on the lines, we may be asking too much of human nature by expecting everyone to do the right thing voluntarily. Many of the greatest public health campaigns in history have necessitated an authoritarian style, and it may be necessary for people all over the world to temporarily embrace and accept new restriction measures they would otherwise justifiably describe as draconian. Perhaps the single most important take-home message of the widely-accepted 80-20 rule of epidemiology (less than 20% of people cause more than 80% of transmission)^110^ is that the extremes of human circumstances and behaviour, especially during mass gatherings and population movements, are more important to the survival of pathogens than the average. It inevitably follows that such exceptions are vitally important to target if one wishes to eliminate COVID-19.^111-114^

Again, it is worth remembering Typhoid Mary^109^, who resisted repeated efforts to get her out of the kitchen and did nothing to disprove stereotypes about the stubborn Irish. She repeatedly returned to working as a cook because it paid better and frequently changed jobs as people fell ill around her, even changing her name to evade more than 30 years of quarantines imposed on her. It took 4 policemen over three hours to apprehend her despite a stealthy approach and forced entry to her home. Eventually Mary was found hiding in an outside closet at the rear of a neighbour’s house, and things remained spicy following her arrest:

“She fought and struggled and cursed. I tried to explain to her that I only wanted the specimens and that then she could go back home. She again refused and I told the policemen to pick her up and put her in the ambulance. This we did and the ride down to the hospital was quite a wild one.”^109^ When we read about the ongoing “Coronavirus challenge” game mediated through social media, we are inclined to think the spirit of Mary Mallon is alive and well and needs to be curbed. The experiences of those who knew Mary Mallon seem extreme but are difficult to disregard completely in the context of a pandemic threatening a global population of over 7 billion people with more eccentric characters, miscreants and outright criminals than we would wish in the circumstances:

“Mary was now about forty-eight years of age and a good deal heavier than she was when she slipped through a kitchen full of servants, jumped the back fence and put up a fight with strong young policemen. She was as strong as ever, but she had lost something of that remarkable energy and activity which had characterized her young days and urged her forward to meet undaunted whatever situation the world presented to her. In these eight years since she was first arrested, she had learned what it was to yield to other wills than her own and to know pain.”^109^

### Contact tracing as an epidemiological surveillance platform, rather than an in intervention per se

As for the full the full, timely intervention package simulated in figure 1 (Compare with figure S1), removing contact tracing from the less rigorous but extended lock down intervention package has only a modest effect on the overall containment trajectory (Figure 4E and F), with only 797 more cases and 3 additional fatalities. Under a the far more extreme conditions of a failed containment campaign, followed by a resurgent, full-blown epidemic, contact tracing becomes a rather pointless exercise, even for targeting clinical disease management. At the peak of the epidemic, when over 4 million new symptomatic cases may occur per week and even mortality rate may outstrip testing capacity (Figure 2), so case confirmation success rates may plummet to below 0.2%.

However, such spectacular containment failures are to be avoided at all costs and this simplified model only accounts for the direct preventative effects of follow up on subsequent transmission, so none of these simulations should be used to in any way imply that contact tracing and isolation should be de-prioritized. In particular, it does not account for the invaluable functions of contact tracing for monitoring and characterizing an epidemic,^1,4,6,22,24,77^ and for understanding the influence of interventions on transmission dynamics. For example, the tracing of transmission to a relatively small number of clusters in Korea, and especially the incrimination of venues like the Shincheonji Church provide invaluable insights that guide more rigorous, effective follow up on lock down measures.^64^ In Ireland, early observations that mean size of close contact clusters had shrunk from 20 to 5 were reported to the public as an encouraging early sign that behavioural interventions were impacting risks of onward transmission. Without such essential detailed information about how transmission persists, as well as the strengths and weakness of ongoing intervention efforts, national containment programmes would be flying blind.

It should be noted, however, that testing, contact tracing and isolation of known contacts is only useful as part of a deliberate containment strategy that keeps an epidemic manageably small, and may be particularly useful for extinguishing the remaining embers of an effectivley-contained epidemic.^41^ While testing is always useful for clinical management of severe cases, once 1% or more of the population has been infected even this important subset of cases alone overwhelms testing capacity (Figure 2B, D, E and F) and the fraction of non-severe cases confirmed plummets to negligible levels. In any case, population-wide testing of mildly symptomatic cases becomes unhelpful as a guide to targeting containment measures: How does one selectively target those at immediate risk when that means everyone? And how would we attempt contract tracing if we allowed the epidemic to grow to tens or hundreds of thousands of new cases each week? Note, however, that the expected failure of contact tracing, and indeed testing generally, is just one more good reason to contain national COVID-19 epidemics before they progress from emergencies (Figures 1 and 4) into outright catastrophes (Figure 2).

### Limitations, caveats and comparisons with other models

Like other recent models of COVID-19, our simplified formulation does not attempt to predict complex indirect effects of the pandemic upon morbidity and mortality from other causes that will be exacerbated by the expected pressures on a health system that is already overstretched.^53^ Nor does it attempt to anticipate the extent of economic and social damage that will arise from different epidemic containment scenarios,^53^ partly because doing so would defeat the purpose of developing a simplified arithmetic formulation.

Despite its limitations as a relatively simple and untested model, the predictions described above are consistent with those of most other process-explicit models using more sophisticated mathematical formulations and specialist software,^25-50^ as well as recent reports of heroic^115,116^ success from China.^8,26,41,117,118^ In fact, perhaps the most useful new lessons to be learned come from a few studies that reach substantively different conclusions based on markedly different underlying assumptions.

Our predictions that contact tracing and isolation will play only a minor role in successful containment contrast with those of others ^119^ which assumed asymptomatic carriage by only 10% of cases or less, and which do not account for the detection dilution effect of similar mild symptoms caused by other common pathogens (Figure 2B, D, F, H).

On the other hand, the predictions presented here appear relatively optimistic when compared with recent reports suggesting viral reproduction rates are higher than generally thought^65,67^ because previous analyses failed to consider the likelihood that large fractions of cases may go undetected^8-13,21^ because they exhibit only mild, non-specific symptoms, if any^1-7,9,12,14-21^ As underlined right at the outset of the global response,^79,120^ the most important remaining question that needs to be answered to reduce the uncertainties of model predictions is the extent of asymptomatic carriage and infectiousness. Learning lessons from other diseases like endemic malaria, which is primarily a chronic illness transmitted by semi-immune adult carriers,^121^ the term *asymptomatic* may well be a misnomer not only because some individuals become infectious before exhibiting symptoms,^1,3,5,16,19,22,23,74^ but also because it is often applied to those who shrug off mild symptoms to get on with their daily lives.^8,122,123^

A particularly important caveat arising from current uncertainty about the role of cryptic carriers is that it also has a major influence on estimation of fatality rate for infections rather than clinical cases. The latest analyses allowing for this phenomenon suggest that fatality rates are may be 10 to 40 times lower per infection than per confirmed case,^63,65^ consistent with our conclusion that the vast majority of cases are never confirmed. While fatality rates are difficult to estimate directly,^124,125^ these modelling analyses support the conclusions of the most controlled empirical epidemiological studies, indicating that the COVID-19 fatality rates may be comfortably below 1%.^21^ The surge of severe cases and fatalities in an uncontained epidemic may therefore peak at a far lower level than those predicted in figure 2. However, much of the variation between fatality rate estimates appears related to geographic differences in health system capacity and burden,^125,126^ so low income countries will be much more vulnerable. Even if the best worst-case scenario proves to be less catastrophic than previously projected,^63,65^ it will nevertheless overwhelm critical care capacity several times over and should be avoided if at all possible.

## Conclusions

The current global health emergency demands immediate, bold, pre-emptive decisions in the absence of unambiguous evidence,^127,128^ based on our best understanding of COVID-19 epidemiology as it stands today.^35,129,130^ The three key sequential actions every country needs to embrace as early and emphatically as possible are *contain, eliminate* and *exclude*. Even when faced with the prospect of lock downs lasting 4 months or more, there is no place for more timid terms like *slow, flatten* or *mitigate* when faced with an epidemic capable of overwhelming ICU capacity hundreds of times over or taking several years of restrictions to slowly burn through an entire population at rates that ICUs can cope with.

And tackling this pandemic will rely overwhelmingly upon widespread understanding and mass participation by the entire global public, rather than just the health professionals and high-level decision makers who will lead the response. Currently, half the world’s population is already under lock down of some kind, meaning vertically enforced and severe restrictions of movement and physical interaction, and the remainder will have to follow if the ongoing COVID-19 virus pandemic is to be contained. Faced with such brutally difficult decisions, it is essential to policy-makers, health professionals and the general public that as many people as possible understand why lock down interventions represent the only realistic way for individual countries to contain their national-level epidemics before they turn into public health catastrophes. It also vital for as many people as possible to understand why these need to be implemented so early, so aggressively and for such extended periods.

Over the medium-to-long term, it will also be vital for us all to understand why widespread national action and international co-operation^117,118^ will be required to conditionally re-open trade and travel between countries that have successfully eliminated local transmission. As explained by the simplified simulations presented here, this appears to be the only means by which national elimination efforts can be sustained, following which pandemic eradication may be pursued at global level.

## Data Availability

All methods and simulation tools described herein are available as supplementary online material and as links.

https://skiware.shinyapps.io/COVID19/

---

**Supplementary Figure 1.**
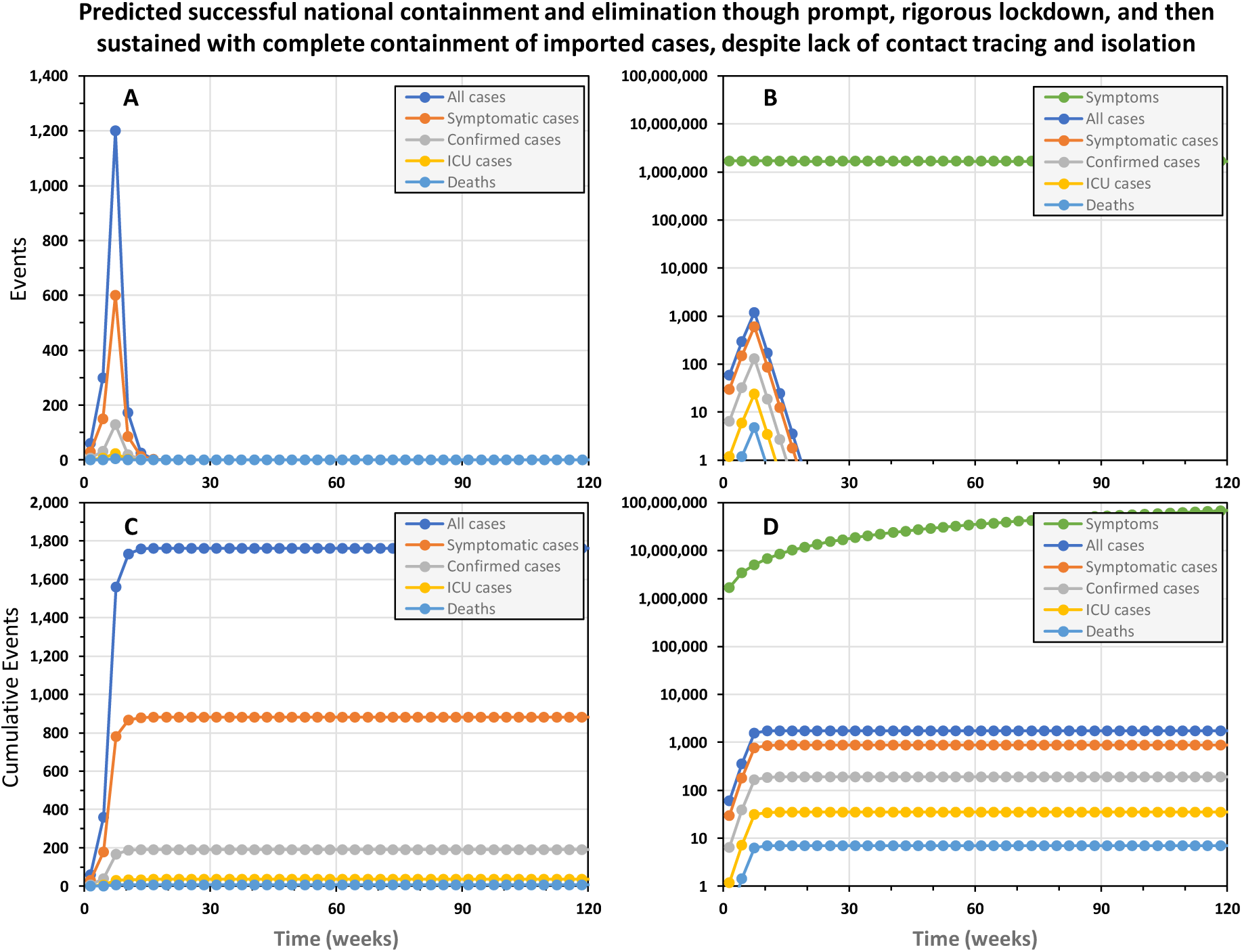
The predicted trajectory of a successfully contained national COVID-19 epidemic in the United Republic of Tanzania, achieved without any contact tracing and isolation. In this simulation, a rigorous 15-week lock down was initiated from week 5 onwards and complemented by complete containment of imported cases, as well as contact tracing and isolation of confirmed cases. Rigorous lock down was assumed to achieve 90% reduction of exposure behaviours by 90% of the population. Complete 100% containment of imported cases assumes that all inbound international visitors are fully isolated for three weeks, ^2,3,5,7,69,70,78^ except those coming from countries that may be certified as free of local transmission by WHO in the future. However, these simulations differ from figure 1 in that absolutely no contact tracing and isolation was assumed.

**Supplementary Figure 2.**
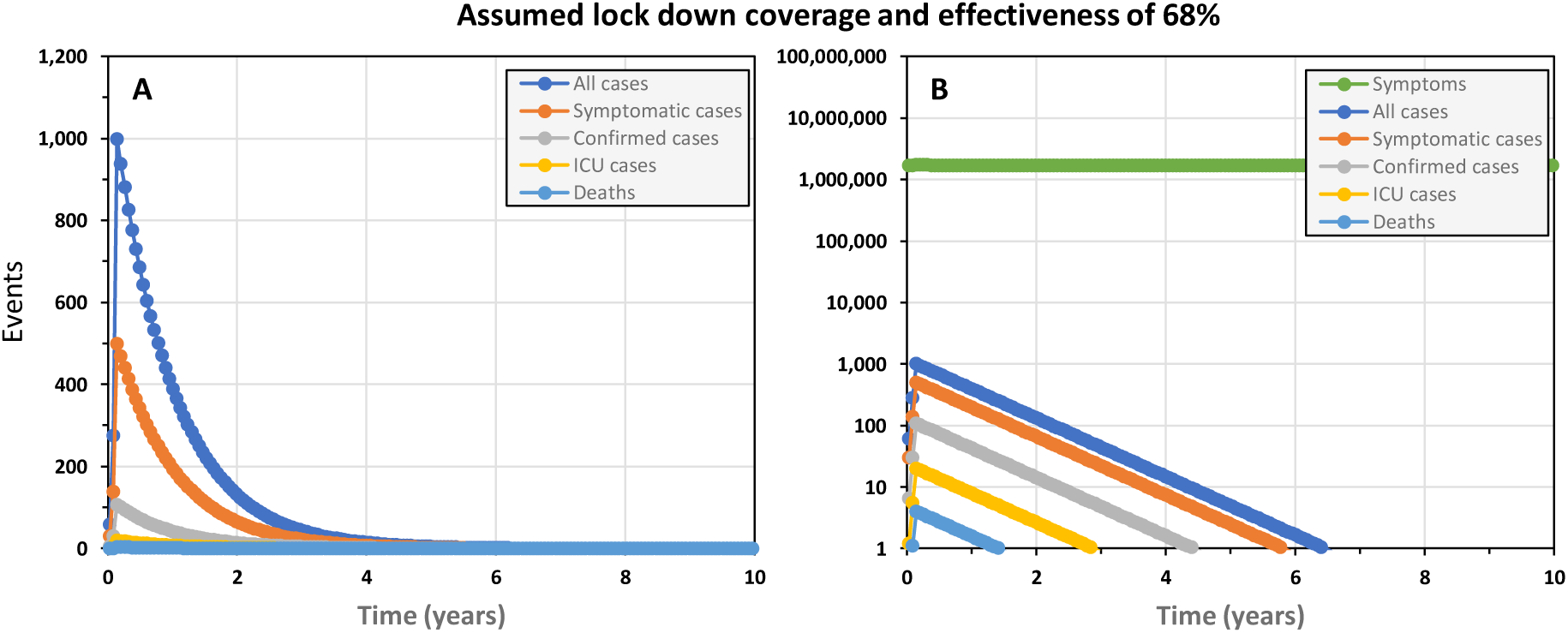
The simulated epidemic trajectory for a less robust national COVID-19 epidemic containment responses in the United Republic of Tanzania than that illustrated in figure 1, intended to *flatten the curve* enough for national health system capacity to cope while also excluding importation of new cases. This simulation has identical input parameters to figure 3E and F except that complete importation containment is assumed.

## Literature Cited

1. Yu P, Zhu J, Zhang Z, Han Y, Huang L. A familial cluster of infection associated with the 2019 novel coronavirus indicating potential person-to-person transmission during the incubation period. J Infect Dis 2020; Electronic pubication ahead of print.

2. Kam KQ, Yung CF, Cui L, et al. A Well Infant with Coronavirus Disease 2019 (COVID-19) with High Viral Load. Clin Infect Dis 2020; Electronic pubication ahead of print.

3. Hu Z, Song C, Xu C, et al. Clinical characteristics of 24 asymptomatic infections with COVID-19 screened among close contacts in Nanjing, China. Sci China Life Sci 2020; Electronic pubication ahead of print.

4. Lu S, Lin J, Zhang Z, et al. Alert for non-respiratory symptoms of Coronavirus Disease 2019 (COVID-19) patients in epidemic period: A case report of familial cluster with three asymptomatic COVID-19 patients. J Med Virol 2020; Electronic pubication ahead of print.

5. Rothe C, Schunk M, Sothmann P, et al. Transmission of 2019-nCoV Infection from an Asymptomatic Contact in Germany. N Engl J Med 2020; 382(10): 970–1.

6. Li C, Ji F, Wang L, et al. Asymptomatic and Human-to-Human Transmission of SARS-CoV-2 in a 2-Family Cluster, Xuzhou, China. Emerg Infect Dis 2020; 26(7) ; Electronic pubication ahead of print.

7. Zou L, Ruan F, Huang M, et al. SARS-CoV-2 Viral Load in Upper Respiratory Specimens of Infected Patients. N Engl J Med 2020; 382: 1177–9.

8. Wang C, Liu L, Hao X, et al. Evolving Epidemiology and Impact of Non-pharmaceutical Interventions on the Outbreak of Coronavirus Disease 2019 in Wuhan, China. MedRXiv 2020.

9. Su L, Ma X, Yu H, et al. The different clinical characteristics of corona virus disease cases between children and their families in China - the character of children with COVID-19. Emerg Microbes Infect 2020; 9(1): 707–13.

10. Nishiura H, Kobayashi T, Suzuki A, et al. Estimation of the asymptomatic ratio of novel coronavirus infections (COVID-19). Int J Infect Dis 2020; Electronic pubication ahead of print.

11. Mizumoto K, Kagaya K, Zarebski A, Chowell G. Estimating the asymptomatic proportion of coronavirus disease 2019 (COVID-19) cases on board the Diamond Princess cruise ship, Yokohama, Japan, 2020. Euro Surveill 2020; 25(10); Electronic pubication ahead of print.

12. Qiu H, Wu J, Hong L, Luo Y, Song Q, Chen D. Clinical and epidemiological features of 36 children with coronavirus disease 2019 (COVID-19) in Zhejiang, China: an observational cohort study. Lancet Infect Dis 2020; Electronic pubication ahead of print.

13. Dong X, Cao YY, Lu XX, et al. Eleven Faces of Coronavirus Disease 2019. Allergy 2020; Electronic pubication ahead of print.

14. Lai CC, Liu YH, Wang CY, et al. Asymptomatic carrier state, acute respiratory disease, and pneumonia due to severe acute respiratory syndrome coronavirus 2 (SARS-CoV-2): Facts and myths. J Microbiol Immunol Infect 2020; Electronic pubication ahead of print.

15. Tang A, Tong ZD, Wang HL, et al. Detection of Novel Coronavirus by RT-PCR in Stool Specimen from Asymptomatic Child, China. Emerg Infect Dis 2020; 26(6); Electronic pubication ahead of print.

16. Covid-19 National Emergency Response Center E, Case Management Team KCfDC, Prevention. Early Epidemiological and Clinical Characteristics of 28 Cases of Coronavirus Disease in South Korea. Osong Public Health Res Perspect 2020; 11(1): 8-14; Electronic pubication ahead of print.

17. Tian S, Hu N, Lou J, et al. Characteristics of COVID-19 infection in Beijing. The Journal of infection 2020; 80(4): 401–6.

18. Shi H, Han X, Jiang N, et al. Radiological findings from 81 patients with COVID-19 pneumonia in Wuhan, China: a descriptive study. Lancet Infect Dis 2020; 20(4): 425–34.

19. Hoehl S, Rabenau H, Berger A, et al. Evidence of SARS-CoV-2 Infection in Returning Travelers from Wuhan, China. N Engl J Med 2020; 382(13): 1278–80.

20. Ki M, Task Force for -nCo V. Epidemiologic characteristics of early cases with 2019 novel coronavirus (2019-nCoV) disease in Korea. Epidemiol Health 2020; 42: e2020007.

21. Nishiura H, Kobayashi T, Yang Y, et al. The Rate of Underascertainment of Novel Coronavirus (2019-nCoV) Infection: Estimation Using Japanese Passengers Data on Evacuation Flights. J Clin Med 2020; 9(2); Electronic pubication ahead of print.

22. Li P, Fu JB, Li KF, et al. Transmission of COVID-19 in the terminal stage of incubation period: a familial cluster. Int J Infect Dis 2020; Electronic pubication ahead of print.

23. Du Z, Xu X, Wu Y, Wang L, Cowling BJ, Meyers LA. Serial Interval of COVID-19 among Publicly Reported Confirmed Cases. Emerg Infect Dis 2020; 26(6); Electronic pubication ahead of print.

24. Ghinai I, McPherson TD, Hunter JC, et al. First known person-to-person transmission of severe acute respiratory syndrome coronavirus 2 (SARS-CoV-2) in the USA. Lancet 2020; Electronic pubication ahead of print.

25. Li R, Pei S, Chen B, et al. Substantial undocumented infection facilitates the rapid dissemination of novel coronavirus (SARS-CoV2). Science 2020; Electronic pubication ahead of print.

26. Prem K, Liu Y, Russell TW, et al. The effect of control strategies to reduce social mixing on outcomes of the COVID-19 epidemic in Wuhan, China: a modelling study. Lancet Public Health 2020; Electronic pubication ahead of print.

27. Zhao S, Chen H. Modeling the epidemic dynamics and control of COVID-19 outbreak in China. Quant Biol 2020: 1–9.

28. Mandal S, Bhatnagar T, Arinaminpathy N, et al. Prudent public health intervention strategies to control the coronavirus disease 2019 transmission in India: A mathematical model-based approach. Indian J Med Res 2020; Electronic pubication ahead of print.

29. Ferguson NM, Laydon D, Nedjati-Gilani G, et al. Impact of non-pharmaceutical interventions (NPIs) to reduce COVID-19 mortality and healthcare demand: WHO Collaborating Centre for Infectious Disease Modelling, MRC Centre for Global Infectious Disease Analysis, Abdul Latif Jameel Institute for Disease and Emergency Analytics & Imperial College London, 2020.

30. Koo JR, Cook AR, Park M, et al. Interventions to mitigate early spread of SARS-CoV-2 in Singapore: a modelling study. Lancet Infect Dis 2020; Electronic pubication ahead of print.

31. Wu JT, Leung K, Leung GM. Nowcasting and forecasting the potential domestic and international spread of the 2019-nCoV outbreak originating in Wuhan, China: a modelling study. Lancet 2020; 395(10225): 689–97.

32. Roosa K, Lee Y, Luo R, et al. Real-time forecasts of the COVID-19 epidemic in China from February 5th to February 24th, 2020. Infect Dis Model 2020; 5: 256–63.

33. Roosa K, Lee Y, Luo R, et al. Short-term Forecasts of the COVID-19 Epidemic in Guangdong and Zhejiang, China: February 13-23, 2020. J Clin Med 2020; 9(2); Electronic pubication ahead of print.

34. Tariq A, Lee Y, Roosa K, Blumberg S, Yan P, Ma S. Real-time monitoring the transmission potential of COVID-19 in Singapore. medRxiv 2020.

35. Anderson RM, Heesterbeek H, Klinkenberg D, Hollingsworth TD. How will country-based mitigation measures influence the course of the COVID-19 epidemic? Lancet 2020; 395(10228): 931–4.

36. Walker PGT, Whittaker C, Watson O, et al. The Global Impact of COVID-19 and Strategies for Mitigation and Suppression: WHO Collaborating Centre for Infectious Disease Modelling, MRC Centre for Global Infectious Disease Analysis, Abdul Latif Jameel Institute for Disease and Emergency Analytics and Imperial College London, 2020.

37. De Salazar PM, Niehus R, Taylor A, Buckee CO, Lipsitch M. Identifying Locations with Possible Undetected Imported Severe Acute Respiratory Syndrome Coronavirus 2 Cases by Using Importation Predictions. Emerg Infect Dis 2020; 26(7); Electronic pubication ahead of print.

38. Karako K, Song P, Chen Y, Tang W. Analysis of COVID-19 infection spread in Japan based on stochastic transition model. Biosci Trends 2020; Electronic pubication ahead of print.

39. Kuniya T. Prediction of the Epidemic Peak of Coronavirus Disease in Japan, 2020. J Clin Med 2020; 9(3); Electronic pubication ahead of print.

40. Neher RA, Dyrdak R, Druelle V, Hodcroft EB, Albert J. Potential impact of seasonal forcing on a SARS-CoV-2 pandemic. Swiss Med Wkly 2020; 150: w20224.

41. Tang B, Xia F, Tang S, et al. The effectiveness of quarantine and isolation determine the trend of the COVID-19 epidemics in the final phase of the current outbreak in China. Int J Infect Dis 2020; Electronic pubication ahead of print.

42. Kucharski AJ, Russell TW, Diamond C, et al. Early dynamics of transmission and control of COVID-19: a mathematical modelling study. Lancet Infect Dis 2020; Electronic pubication ahead of print.

43. Choi SC, Ki M. Estimating the reproductive number and the outbreak size of Novel Coronavirus disease (COVID-19) using mathematical model in REpublic of Korea. Epidemiol Health 2020: e2020011.

44. Lin Q, Zhao S, Gao D, et al. A conceptual model for the coronavirus disease 2019 (COVID-19) outbreak in Wuhan, China with individual reaction and governmental action. Int J Infect Dis 2020; 93: 211–6.

45. Chinazzi M, Davis JT, Ajelli M, et al. The effect of travel restrictions on the spread of the 2019 novel coronavirus (COVID-19) outbreak. Science 2020; Electronic pubication ahead of print.

46. Fang Y, Nie Y, Penny M. Transmission dynamics of the COVID-19 outbreak and effectiveness of government interventions: A data-driven analysis. J Med Virol 2020; Electronic pubication ahead of print.

47. Wang H, Wang Z, Dong Y, et al. Phase-adjusted estimation of the number of Coronavirus Disease 2019 cases in Wuhan, China. Cell Discov 2020; 6: 10.

48. Boldog P, Tekeli T, Vizi Z, Denes A, Bartha FA, Rost G. Risk Assessment of Novel Coronavirus COVID-19 Outbreaks Outside China. J Clin Med 2020; 9(2); Electronic pubication ahead of print.

49. Tang B, Bragazzi NL, Li Q, Tang S, Xiao Y, Wu J. An updated estimation of the risk of transmission of the novel coronavirus (2019-nCov). Infect Dis Model 2020; 5: 248–55.

50. Tang B, Wang X, Li Q, et al. Estimation of the Transmission Risk of the 2019-nCoV and Its Implication for Public Health Interventions. J Clin Med 2020; 9(2); Electronic pubication ahead of print.

51. Briggs DJ, Sabel CE, Lee K. Uncertainty in epidemiology and health risk and impact assessment. Environ Geochem Health 2009; 31(2): 189–203.

52. Christley RM, Mort M, Wynne B, et al. “Wrong, but useful”: negotiating uncertainty in infectious disease modelling. PLoS One 2013; 8(10): e76277.

53. Enserink M, Kupferschmidt K. With COVID-19, modeling takes on life and death importance. Science 2020; 367(6485): 1414–5.

54. Haider N, Yavlinsky A, Simons D, et al. Passengers’ destinations from China: low risk of Novel Coronavirus (2019-nCoV) transmission into Africa and South America. Epidemiol Infect 2020; 148: e41.

55. Gilbert M, Pullano G, Pinotti F, et al. Preparedness and vulnerability of African countries against importations of COVID-19: a modelling study. Lancet 2020; 395(10227): 871–7.

56. Nkengasong JN, Mankoula W. Looming threat of COVID-19 infection in Africa: act collectively, and fast. Lancet 2020; 395(10227): 841–2.

57. Agyeman AA, Laar A, Ofori-Asenso R. Will COVID-19 be a litmus test for post-Ebola sub-Saharan Africa? J Med Virol 2020; Electronic pubication ahead of print.

58. Makoni M. Africa prepares for coronavirus. Lancet 2020; 395(10223): 483.

59. Lloyd-Sherlock P, Ebrahim S, Geffen L, McKee M. Bearing the brunt of covid-19: older people in low and middle income countries. BMJ 2020; 368: m1052.

60. Wang J, Xu C, Wong YK, et al. Preparedness is essential for malaria-endemic regions during the COVID-19 pandemic. Lancet 2020; Electronic pubication ahead of print.

61. Engdahl Mtango S, Lugazia E, Baker U, Johansson Y, Baker T. Referral and admission to intensive care: A qualitative study of doctors’ practices in a Tanzanian university hospital. PLoS One 2019; 14(10): e0224355.

62. Pearson CAB, Van Schalkwyk C, Foss A, O’Reilly K, Pulliam J. Projection of early spread of COVID19 in Africa as of 25 March 2020: London School of Hygiene & Tropical Medicine, 2020.

63. Anastassopoulou C, Russo L, Tsakris A, Siettos C. Data-based analysis, modelling and forecasting of the COVID-19 outbreak. PLoS One 2020; 15(3): e0230405.

64. Shim E, Tariq A, Choi W, Lee Y, Chowell G. Transmission potential and severity of COVID-19 in South Korea. Int J Infect Dis 2020; Electronic pubication ahead of print.

65. Mizumoto K, Kagaya K, Chowell G. Early epidemiological assessment of the transmission potential and virulence of coronavirus disease 2019 (COVID-19) in Wuhan City: China, January-February, 2020. MedRXiv 2020.

66. Read JM, Bridgen JR, Cummings DA, Ho A, Jewell CP. Novel coronavirus 2019-nCoV: Early estimation of epidemiological parameters and epidemic predictions. MedRXiv 2020.

67. Sanche S, Lin YT, Xu C, Romero-Severson E, Hengartner N, Ke R. The Novel Coronavirus, 2019-nCoV, is Highly Contagious and More Infectious Than Initially Estimated. MedRXiv 2020.

68. Chen TM, Rui J, Wang QP, Zhao ZY, Cui JA, Yin L. A mathematical model for simulating the phase-based transmissibility of a novel coronavirus. Infect Dis Poverty 2020; 9(1): 24.

69. Linton NM, Kobayashi T, Yang Y, et al. Incubation Period and Other Epidemiological Characteristics of 2019 Novel Coronavirus Infections with Right Truncation: A Statistical Analysis of Publicly Available Case Data. J Clin Med 2020; 9(2) ; Electronic pubication ahead of print.

70. Xing Y, Mo P, Xiao Y, Zhao O, Zhang Y, Wang F. Post-discharge surveillance and positive virus detection in two medical staff recovered from coronavirus disease 2019 (COVID-19), China, January to February 2020. Euro Surveill 2020; 25(10); Electronic pubication ahead of print.

71. National Bureau of Statistics & Ministry of Finance and Planning Dar es Salaam and the Office of the Chief Government Statistician & Ministry of Finance and Planning Zanzibar. National Population Projections: United R Epublic of Tanzania, 2018.

72. BMJ Best Practice. Common Cold. Br Med J 2020.

73. Gostic K, Gomez AC, Mummah RO, Kucharski AJ, Lloyd-Smith JO. Estimated effectiveness of symptom and risk screening to prevent the spread of COVID-19. eLife 2020; 9; Electronic pubication ahead of print.

74. Nishiura H, Linton NM, Akhmetzhanov AR. Serial interval of novel coronavirus (COVID-19) infections. Int J Infect Dis 2020; 93: 284–6.

75. Guan WJ, Ni ZY, Hu Y, et al. Clinical Characteristics of Coronavirus Disease 2019 in China. N Engl J Med 2020; Electronic pubication ahead of print.

76. Yang S, Cao P, Du P, et al. Early estimation of the case fatality rate of COVID-19 in mainland China: a data-driven analysis. Ann Transl Med 2020; 8(4): 128.

77. Pung R, Chiew CJ, Young BE, et al. Investigation of three clusters of COVID-19 in Singapore: implications for surveillance and response measures. Lancet 2020; 395(10229): 1039–46.

78. Ling Y, Xu SB, Lin YX, et al. Persistence and clearance of viral RNA in 2019 novel coronavirus disease rehabilitation patients. Chin Med J (Engl) 2020; Electronic pubication ahead of print.

79. Li Q, Guan X, Wu P, et al. Early Transmission Dynamics in Wuhan, China, of Novel Coronavirus-Infected Pneumonia. N Engl J Med 2020; Electronic pubication ahead of print.

80. Ministry of Health, Community Development, Gender, Elderly and Children. Health Statistical Tables and Figures for 2017: United R Epublic of Tanzania, 2018.

81. Ngowi AF, Kamazima SR, Kibusi S, Gesase A, Bali T. Women’s determinant factors for preferred place of delivery in Dodoma region Tanzania: a cross sectional study. Reprod Health 2017; 14(1): 112.

82. Lyimo MA, Mosha IH. Reasons for delay in seeking treatment among women with obstetric fistula in Tanzania: a qualitative study. BMC Womens Health 2019; 19(1): 93.

83. Nyamuryekung’e KK, Lahti S, Tuominen R. Costs of dental care and its financial impacts on patients in a population with low availability of services. Community Dent Health 2019; 36(2): 131–6.

84. Mhalu G, Hella J, Mhimbira F, et al. Pathways and associated costs of care in patients with confirmed and presumptive tuberculosis in Tanzania: A cross-sectional study. BMJ Open 2019; 9(4): e025079.

85. Bryson-Cahn C, Duchin J, Makarewicz VA, et al. A Novel Approach for a Novel Pathogen: using a home assessment team to evaluate patients for 2019 novel coronavirus (SARS-CoV-2). Clin Infect Dis 2020; Electronic pubication ahead of print.

86. Glauser W. Proposed protocol to keep COVID-19 out of hospitals. CMAJ 2020; 192(10): E264–E5.

87. Mahase E. Coronavirus: home testing pilot launched in London to cut hospital visits and ambulance use. BMJ 2020; 368: m621.

88. Mahase E. Coronavirus: Wales tests 90% of suspected patients in their own home. BMJ 2020; 368: m698.

89. Ministry of Health, Community Development, Gender, Elderly and Children. Mid-Term Review of the Health Sector Strategic Plan IV for 2015 - 2020: Main Report: United R Epublic of Tanzania, 2019.

90. Baynes C, Semu H, Baraka J, et al. An exploration of the feasibility, acceptability, and effectiveness of professional, multitasked community health workers in Tanzania. Glob Public Health 2017; 12(8): 1018–32.

91. Baynes C, Mboya D, Likasi S, et al. Quality of Sick Child-Care Delivered by Community Health Workers in Tanzania. Int J Health Policy Manag 2018; 7(12): 1097–109.

92. Ngilangwa DP, Mgomella GS. Factors associated with retention of community health workers in maternal, newborn and child health programme in Simiyu Region, Tanzania. Afr J Prim Health Care Fam Med 2018; 10(1): e1–e8.

93. Stresman G, Cameron A, Drakeley C. Freedom from Infection: Confirming Interruption of Malaria Transmission. Trends Parasitol 2017; 33(5): 345–52.

94. Cameron AR, Baldock FC. Two-stage sampling in surveys to substantiate freedom from disease. Prev Vet Med 1998; 34(1): 19–30.

95. Cameron AR, Baldock FC. A new probability formula for surveys to substantiate freedom from disease. Prev Vet Med 1998; 34(1): 1–17.

96. Martin PA, Cameron AR, Barfod K, Sergeant ES, Greiner M. Demonstrating freedom from disease using multiple complex data sources 2: case study--classical swine fever in Denmark. Prev Vet Med 2007; 79(2-4): 98–115.

97. Martin PA, Cameron AR, Greiner M. Demonstrating freedom from disease using multiple complex data sources 1: a new methodology based on scenario trees. Prev Vet Med 2007; 79(2-4): 71–97.

98. Cameron A, Njeumi F, Chibeu D, Martin T. Risk-based disease surveillance: A manual for veterinarians on the design and analysis of surveillance for demonstration of freedom from disease: Food and Agriculture Organization of the United Nations; 2014.

99. National Bureau of Statistics. Tanzania in Figures. Dodoma: United REpublic of Tanzania, 2018.

100. Killeen GF, Seyoum A, Sikaala CH, et al. Eliminating malaria vectors. Parasit Vectors 2013; 6: 172.

101. Ahmed F, Ahmed N, Pissarides C, Stiglitz J. Why inequality could spread COVID-19. Lancet Public Health 2020; Electronic pubication ahead of print.

102. Poole DN, Escudero DJ, Gostin LO, Leblang D, Talbot EA. Responding to the COVID-19 pandemic in complex humanitarian crises. Int J Equity Health 2020; 19(1): 41.

103. Liem A, Wang C, Wariyanti Y, Latkin CA, Hall BJ. The neglected health of international migrant workers in the COVID-19 epidemic. Lancet Psychiatry 2020; 7(4): e20.

104. Zhang Z, Liu S, Xiang M, et al. Protecting healthcare personnel from 2019-nCoV infection risks: lessons and suggestions. Front Med 2020; Electronic pubication ahead of print.

105. Rose C. Am I Part of the Cure or Am I Part of the Disease? Keeping Coronavirus Out When a Doctor Comes Home. N Engl J Med 2020; Electronic pubication ahead of print.

106. Bedford J, Enria D, Giesecke J, et al. COVID-19: towards controlling of a pandemic. Lancet 2020; 395(10229): 1015–8.

107. Bhadelia N. Coronavirus: hospitals must learn from past pandemics. Nature 2020; 578(7794): 193.

108. Mizumoto K, Chowell G. Transmission potential of the novel coronavirus (COVID-19) onboard the diamond Princess Cruises Ship, 2020. Infect Dis Model 2020; 5: 264–70.

109. Soper GA. The Curious Career of Typhoid Mary. Bull N Y Acad Med 1939; 15(10): 698–712.

110. Woolhouse MEJ, Dye C, Etard JF, et al. Heterogeneities in the transmission of infectious agents: implications for the design of control programs. Proc Natl Acad Sci USA 1997; 94: 338–42.

111. Chen S, Yang J, Yang W, Wang C, Barnighausen T. COVID-19 control in China during mass population movements at New Year. Lancet 2020; 395(10226): 764–6.

112. Frieden TR, Lee CT. Identifying and Interrupting Superspreading Events-Implications for Control of Severe Acute Respiratory Syndrome Coronavirus 2. Emerg Infect Dis 2020; 26(6); Electronic pubication ahead of print.

113. Ebrahim SH, Memish ZA. COVID-19: preparing for superspreader potential among Umrah pilgrims to Saudi Arabia. Lancet 2020; 395(10227): e48.

114. Liu Y, Eggo RM, Kucharski AJ. Secondary attack rate and superspreading events for SARS-CoV-2. Lancet 2020; 395(10227): e47.

115. Green A. Li Wenliang. Lancet 2020; 395: 682.

116. Calisher C, Carroll D, Colwell R, et al. Statement in support of the scientists, public health professionals, and medical professionals of China combatting COVID-19. Lancet 2020; 395(10226): e42–e3.

117. Zhang S, Wang Z, Chang R, et al. COVID-19 containment: China provides important lessons for global response. Front Med 2020; Electronic pubication ahead of print.

118. Gong F, Xiong Y, Xiao J, et al. China’s local governments are combating COVID-19 with unprecedented responses - from a Wenzhou governance perspective. Front Med 2020; Electronic pubication ahead of print.

119. Hellewell J, Abbott S, Gimma A, et al. Feasibility of controlling COVID-19 outbreaks by isolation of cases and contacts. Lancet Glob Health 2020; 8(4): e488–e96.

120. Munster VJ, Koopmans M, van Doremalen N, van Riel D, de Wit E. A Novel Coronavirus Emerging in China - Key Questions for Impact Assessment. N Engl J Med 2020; 382(8): 692–4.

121. Ross A, Killeen GF, Smith TA. Relationships of host infectivity to mosquitoes and asexual parasite density in Plasmodium falciparum. Am J Trop Med Hyg 2006; 75 (Suppl. 2): 32–7.

122. Chen I, Clarke SE, Gosling R, et al. “Asymptomatic” malaria: A chronic and debilitating infection that should be treated. PLoS Med 2016; 13: e1001942.

123. Sifft KC, Geus D, Mukampunga C, et al. Asymptomatic only at first sight: malaria infection among schoolchildren in highland Rwanda. Malar J 2016; 15(1): 553.

124. Kobayashi T, Jung SM, Linton NM, et al. Communicating the Risk of Death from Novel Coronavirus Disease (COVID-19). J Clin Med 2020; 9(2); Electronic pubication ahead of print.

125. Battegay M, Kuehl R, Tschudin-Sutter S, Hirsch HH, Widmer AF, Neher RA. 2019-novel Coronavirus (2019-nCoV): estimating the case fatality rate - a word of caution. Swiss Med Wkly 2020; 150: w20203.

126. Ji Y, Ma Z, Peppelenbosch MP, Pan Q. Potential association between COVID-19 mortality and health-care resource availability. Lancet Glob Health 2020; 8(4): e480.

127. Smith GC, Pell JP. Parachute use to prevent death and major trauma related to gravitational challenge: systematic review of randomised controlled trials. BMJ 2003; 327(7429): 1459–61.

128. Horton R. Offline: Apostasy against the public health elites. Lancet 2018; 391: 643.

129. Fauci AS, Lane HC, Redfield RR. Covid-19 - Navigating the Uncharted. N Engl J Med 2020; 382(13): 1268–9.

130. Horton R. Offline: COVID-19 and the NHS-”a national scandal”. Lancet 2020; 395(10229): 1022.

